# Exploring factors influencing user perspective of ChatGPT as a technology that assists in healthcare decision making: A cross sectional survey study

**DOI:** 10.1101/2023.12.07.23299685

**Authors:** Avishek Choudhury, Safa Elkefi, Achraf Tounsi

**Author notes:** **Corresponding:** Avishek Choudhury (Ph.D.) Assistant Professor Industrial and Management Systems Engineering, Benjamin M. Statler College of Engineering and Mineral Resources, West Virginia University, 1306 Evansdale Drive Morgantown, West Virginia 26506 USA.

## Abstract

As ChatGPT emerges as a potential ally in healthcare decision-making, it is imperative to investigate how users leverage and perceive it. The repurposing of technology is innovative but brings risks, especially since AI’s effectiveness depends on the data it’s fed. In healthcare, where accuracy is critical, ChatGPT might provide sound advice based on current medical knowledge, which could turn into misinformation if its data sources later include erroneous information. Our study assesses user perceptions of ChatGPT, particularly of those who used ChatGPT for healthcare-related queries. By examining factors such as competence, reliability, transparency, trustworthiness, security, and persuasiveness of ChatGPT, the research aimed to understand how users rely on ChatGPT for health-related decision-making. A web-based survey was distributed to U.S. adults using ChatGPT at least once a month. Data was collected from February to March 2023. Bayesian Linear Regression was used to understand how much ChatGPT aids in informed decision-making. This analysis was conducted on subsets of respondents, both those who used ChatGPT for healthcare decisions and those who did not. Qualitative data from open-ended questions were analyzed using content analysis, with thematic coding to extract public opinions on urban environmental policies. The coding process was validated through inter-coder reliability assessments, achieving a Cohen’s Kappa coefficient of 0.75. Six hundred and seven individuals responded to the survey. Respondents were distributed across 306 US cities of which 20 participants were from rural cities. Of all the respondents, 44 used ChatGPT for health-related queries and decision-making. While all users valued the content quality, privacy, and trustworthiness of ChatGPT across different contexts, those using it for healthcare information place a greater emphasis on safety, trust, and the depth of information. Conversely, users engaging with ChatGPT for non-healthcare purposes prioritize usability, human-like interaction, and unbiased content. In conclusion our study findings suggest a clear demarcation in user expectations and requirements from AI systems based on the context of their use.

## Introduction

In the discourse on the emergence and integration of artificial intelligence (AI) in daily life, the rise of generative pre-trained transformers (GPT) like ChatGPT stands as a hallmark of innovation. As an AI model developed by OpenAI, ChatGPT has garnered widespread attention and adoption for its ability to generate human-like text, engaging in conversations and answering queries with a semblance of understanding previously reserved for human intellect.

The application of ChatGPT extends well beyond its initial conception, echoing a common narrative in the evolution of technology where tools are repurposed in manners unforeseen by their creators. ChatGPT, while intended for conversational assistance, has been appropriated for diverse purposes, from drafting legal and academic documents to creating artistic compositions [1–5]. The recontextualization of technology, while innovative, surfaces inherent risks. As with any tool, the efficacy of AI is contingent upon the parameters of its operation—parameters that are, in the case of AI like ChatGPT, defined by data [6]. Such dependencies on data inputs introduces a temporal dimension to its reliability. The AI’s performance today may not be indicative of its performance in the future. The temporal variability is crucial when considering ChatGPT’s role in domains where accuracy is vital, such as healthcare. Just as a treatment’s efficacy may change over time with new medical discoveries, ChatGPT’s responses are subject to the ebb and flow of the data it consumes. A user may receive sound medical advice one month, only to be misinformed the next, should the AI’s data sources become tainted with erroneous information [7].

User trust in technology is often built over time through consistent and reliable performance [8]. However, in the context of AI, this trust may become a liability if users become complacent, overlooking the potential for AI responses to degrade as the data landscape shifts. User trust in such AI and the extent to which they perceive it to be a helpful decision-making assistant depends on multiple factors such as socio-ethical considerations, technical and design features, user characteristics, and expertise [9, 10]. When users are well-versed in the mechanics of ChatGPT and the principles guiding its responses, they can navigate its capabilities with discernment, appropriately integrating it into their decision-making processes. Conversely, misunderstanding of ChatGPT’s functioning can result in hyped expectations and distorted perception of the technology, leading to unwanted consequences when leveraged for critical applications like healthcare. In other words, if the user is not qualified to validate ChatGPT’s response, the risk or probability of decision errors increases substantially. Furthermore, Trust in AI can also be influenced by personal, organizational, and policy factors [7], as well as properties of the AI system, including controllability, model complexity, embedded biases, and reliability [11]. Transparency is often viewed as a prerequisite for trust in society, and the positive correlation between AI system transparency and trust has been confirmed by previous empirical studies [12, 13].

ChatGPT’s ability to deliver information persuasively also determines how and to what extent people use it. While a convincing articulation can enhance user confidence in the AI’s suggestions, it must be carefully calibrated with the accuracy of the content provided. Persuasiveness without the foundation of reliable and accurate information can lead to misplaced trust and potential misjudgments, especially in high-stakes scenarios such as healthcare decision-making.

The convergence of reliable performance and security protocols also consolidates user trust. On December 1, 2023, Google noted a critical flaw in ChatGPT highlighting the possibility of breaching its training data [14]. GPT models, including ChatGPT, tend to memorize training data, which can lead to privacy concerns [15]. This is particularly concerning if AI models are trained with personal information, as it could lead to the exposure of sensitive data. While efforts are made to align AI models like ChatGPT to prevent the release of large amounts of training data, there is still a risk of data breaches through targeted attacks.

As ChatGPT emerges as a potential ally in healthcare decision-making, it is imperative to investigate how users leverage and perceive it. Our study assesses user perceptions of ChatGPT, particularly of those who used ChatGPT for healthcare-related queries. By examining factors such as competence, reliability, transparency, trustworthiness, security, and persuasiveness of ChatGPT, the research aimed to understand how users rely on ChatGPT for health-related decision-making. Additionally, this study incorporates content analysis explore user concerns with ChatGPT. Our objective is particularly important in the broader context of AI integration into healthcare systems. As AI technologies like ChatGPT become more prevalent, understanding user perspectives on their effectiveness, trustworthiness, and security becomes crucial. These insights are essential for developing AI tools that are not only technically competent but also aligned with the expectations and concerns of end-users, particularly in sensitive areas such as healthcare.

## Methods

### Ethics statement

The study, bearing the Institutional Review Board (IRB) protocol number 2302725983 and classified as a flex protocol type, received approval from West Virginia University. No identifiers were collected during the study. In compliance with ethical research practices, informed consent was obtained from all participants before initiating the survey. Attached to the survey was a comprehensive cover letter outlining the purpose of the study, the procedure involved, the approximate time to complete the survey, and assurances of anonymity and confidentiality. It also emphasized that participation was completely voluntary, and participants could withdraw at any time without any consequences. The cover letter also included contact information of the researchers for any questions or concerns the participants might have regarding the study. Participants were asked to read through this information carefully and were instructed to proceed with the survey only if they understood and agreed to the terms described, effectively providing their consent to participate in the study.

### Data collection

We distributed a web-based semi structured survey to adults in the United States who actively use ChatGPT at least once a month. We collected the data from February 1^st^, 2023, through March 30^th^, 2023. We conducted a soft launch of the survey and collected 40 responses. A soft launch is a small-scale test of a survey before it is distributed to a larger audience. This soft launch aimed to identify any potential issues with the survey, such as unclear or confusing questions, technical glitches, or other problems that may affect the quality of the data collected. The survey was then distributed to a larger audience across the US.

### Instrument

The survey was designed on Qualtrics and was distributed by Centiment, an paid audience-paneling service to reach a broader population [16]. The survey consisted of 17 Likert scale questions and two open ended questions as reported in Table 1. To ensure response quality, we included a checking question “We would like to ensure you are reading each question and responding thoughtfully. Please select "Green" as your answer.” Questions assessing competence, reliability, transparency, trustworthiness, and integrity of ChatGPT were informed by constructs in technology acceptance models and trust research in information systems.

**Table 1.** Survey questions.

| Questions | Variable name |
| --- | --- |
| How frequently do you use ChatGPT | Use frequency |
| What is the main reason you use ChatGPT? | Use purpose |
| **To what extent do you agree or disagree with the following:** |  |
| ChatGPT is competent in providing the information and guidance. | Competent |
| ChatGPT is reliable in providing consistent and dependable information. | Reliable |
| ChatGPT is transparent. | Transparent |
| ChatGPT is trustworthy. | Trustworthy |
| ChatGPT will not manipulate its responses. | Not manipulate |
| ChatGPT is secure and protects my privacy and confidential information | Secure |
| I am willing to use ChatGPT for healthcare-related queries. | ChatGPT for healthcare queries |
| Benefits of using ChatGPT outweigh any potential risks. | Benefits outweigh risks |
| ChatGPT helps me make informed and timely decisions. | Help make decisions |
| I am willing to take decisions based on the recommendations provided by ChatGPT. | Willing to take decisions |
| I am satisfied with ChatGPT. | Satisfaction |
| I am willing to use ChatGPT in the future. | Intent to use |
| ChatGPT can replace human-to-human interaction. | Replace human interaction |
| ChatGPT is persuasive. | Persuasive |
| What is the highest degree or level of education you have completed? | Education |
| *Describe in your own words, the improvements you would like to see in ChatGPT. | Not applicable |
| *Highlight your concerns with ChatGPT. | Not applicable |
| *\*Open ended questions* |  |

### Statistical analyses

The analysis was divided into quantitative and qualitative. The quantitative analyses involved all the Likert scale variables. We first calculated the data’s descriptive statistics and reported the data’s central tendency, dispersion, and inference.

We then focused on the dataset corresponding to those who used ChatGPT for making healthcare-related decisions. We calculated the multivariate and pairwise normality of the subset of the data. Given the data distribution and normality violation, we conducted the Bayesian correlation test [17, 18]. In the Bayesian Pearson correlation analyses, the strength and direction of the relationships between various perceived attributes of ChatGPT are quantified using Pearson’s correlation coefficient (r) quantifying the evidence for a correlation against the null hypothesis of no correlation. The Bayesian Linear Regression with Jeffreys-Zellner-Siow (JZS) priors for coefficients was conducted to ascertain the extent to which respondents agree that ChatGPT helps them make informed and timely decisions, denoted as "Help make decisions" [19]. The JZS prior is a default prior representing a compromise between informativeness and non-informativeness, providing a reference analysis less sensitive to the prior choice [19]. The analysis was predicated on a uniform prior distribution, reflecting the absence of a priori preferences or knowledge about the importance of the predictors. This non-informative prior ensures that the posterior distributions are primarily influenced by the data rather than by subjective prior beliefs [19, 20]. Model comparison was achieved by contrasting each model with the best-performing model as a reference. The Bayes Factor (BF_10_) was used to quantify the evidence for each model against the best model, and the coefficient of determination (R²) was calculated to assess the proportion of variance explained by the models. BF_10_ provides a measure of evidence of alternative hypothesis (H_1_) over null hypothesis (H_0_), where H_0_ suggests that there is no effect or no difference between the variables being studied, and H_1_ suggests that there is an effect or a difference [21]. Lastly, the posterior distribution of the regression coefficients was summarized, showing the mean, standard deviation (SD), and 95% credible intervals for each factor.

We repeated the same analysis on a subset of our data corresponding to respondents who did not use ChatGPT for health-related decision-making.

The open-ended qualitative data was analyzed using content analysis [22]. The question was designed to gather insights on public opinions regarding urban environmental policies. First the unit of analysis was identified as thematic phrases within each response followed by the development of a coding scheme to categorize the survey responses. The primary coding was conducted by one researcher, with periodic validation checks by a second researcher to maintain coding consistency. All coders engaged in iterative discussions to refine and finalize a set of consensus codes, ensuring that the identified themes accurately captured the essence of the participants’ experiences and perspectives. Inter-coder reliability was assessed using Cohen’s Kappa, yielding a coefficient of 0.75, indicative of substantial agreement [23].

## Result

### Data description and understanding

Six hundred and seven individuals responded to the survey. As illustrated in Fig 1, respondents were distributed across 306 US cities. Only 20 participants were from rural cities. Of all the respondents, 44 used ChatGPT for health-related queries. Other uses of the technology were information gathering (n=219), entertainment (n=203), and problem-solving (n=135), and fun activities (n=6). Note:

**Figure 1.** The figure shows the geographic distribution of study participants from 306 cities across the US. The blue circle size corresponds to the number of responses from each location. The data reveals a high concentration of participants in urban areas across the Eastern Seaboard, parts of the Midwest, and the West Coast, particularly in California, with sparser distribution in the central United States and rural regions.

Table 2 shows the descriptive statistics of study variables. The statistics include the mean, standard error of the mean, a 95% confidence interval for the mean (with upper and lower bounds), standard deviation, Shapiro-Wilk test results, p-value of the Shapiro-Wilk test for normality, and the minimum and maximum values observed for each variable. Perceptions of ChatGPT’s attributes such as competence, reliability, transparency, trustworthiness, and security were rated positively, with all means exceeding 3.0. The Shapiro-Wilk test uniformly indicated non-normality in the data distributions (*p < .001*), suggesting the presence of skewness or kurtosis across variables.

**Table 2.** Descriptive statistics.

|  | Mean | Std.<br>error of<br>mean | 95%<br>confidence<br>interval mean |  | Std.<br>Deviation | Shapiro-Wilk | Min | Max |
| --- | --- | --- | --- | --- | --- | --- | --- | --- |
|  |  |  | Upper | Lower |  |  |  |  |
| Competent | 3.199 | 0.034 | 3.265 | 3.134 | 0.825 | 0.794*** | 1 | 4 |
| Reliable | 3.161 | 0.032 | 3.225 | 3.098 | 0.800 | 0.812*** | 1 | 4 |
| Transparent | 3.119 | 0.035 | 3.187 | 3.050 | 0.859 | 0.817*** | 1 | 4 |
| Trustworthy | 3.168 | 0.034 | 3.235 | 3.101 | 0.837 | 0.807*** | 1 | 4 |
| Not manipulate | 3.099 | 0.036 | 3.169 | 3.029 | 0.881 | 0.823*** | 1 | 4 |
| Secure | 3.273 | 0.033 | 3.338 | 3.209 | 0.811 | 0.775*** | 1 | 4 |
| ChatGPT for healthcare queries | 3.097 | 0.035 | 3.165 | 3.029 | 0.855 | 0.826*** | 1 | 4 |
| Benefits outweigh risks | 3.204 | 0.033 | 3.268 | 3.140 | 0.801 | 0.798*** | 1 | 4 |
| Helps make decisions | 3.257 | 0.032 | 3.320 | 3.194 | 0.790 | 0.782*** | 1 | 4 |
| Satisfaction | 3.244 | 0.031 | 3.305 | 3.183 | 0.764 | 0.792*** | 1 | 4 |
| Willing to take decisions | 3.130 | 0.033 | 3.195 | 3.065 | 0.815 | 0.818*** | 1 | 4 |
| Intent to use | 3.379 | 0.031 | 3.439 | 3.319 | 0.757 | 0.751*** | 1 | 4 |
| Replace human interaction | 2.292 | 0.037 | 3.001 | 2.857 | 0.901 | 0.847*** | 1 | 4 |
| Persuasive | 2.979 | 0.033 | 3.044 | 2.913 | 0.819 | 0.846*** | 1 | 4 |
| Education | 2.855 | 0.036 | 2.925 | 2.785 | 0.884 | 0.873*** | 1 | 5 |

\*\*\* $p < 0.001$

**Table 3.** The Bayesian Pearson Correlation of subset participants who use ChatGPT for health-related queries (n=44). The table details the correlation coefficients (Pearson’s r) among various perceived attributes of ChatGPT.

|  | A | B | C | D | E | F | G | H | I | J | K | L | M | N |
| --- | --- | --- | --- | --- | --- | --- | --- | --- | --- | --- | --- | --- | --- | --- |
| Competent (A) |  |  |  |  |  |  |  |  |  |  |  |  |  |  |
| Reliable (B) | 0.116 |  |  |  |  |  |  |  |  |  |  |  |  |  |
| Transparent (C) | 0.469* | 0.119 |  |  |  |  |  |  |  |  |  |  |  |  |
| Trustworthy (D) | 0.436* | 0.169 | 0.623*** |  |  |  |  |  |  |  |  |  |  |  |
| Not manipulate (E) | 0.342 | 0.174 | 0.496** | 0.322 |  |  |  |  |  |  |  |  |  |  |
| Secure (F) | 0.443* | 0.287 | 0.518** | 0.425* | 0.583*** |  |  |  |  |  |  |  |  |  |
| ChatGPT for healthcare queries (G) | 0.289 | 0.016 | 0.437* | 0.463* | 0.481** | 0.297 |  |  |  |  |  |  |  |  |
| Benefits outweigh risks (H) | 0.415 | 0.132 | 0.287 | 0.274 | 0.384 | 0.390 | 0.532*** |  |  |  |  |  |  |  |
| Helps make decisions (I) | 0.513* | 0.084 | 0.467* | 0.682*** | 0.446* | 0.453* | 0.538*** | 0.430* |  |  |  |  |  |  |
| Satisfaction (J) | 0.367 | 0.074 | 0.498* | 0.467* | 0.285 | 0.569*** | 0.354 | 0.355 | 0.359 |  |  |  |  |  |
| Willing to take decisions (K) | 0.382 | 0.160 | 0.317 | 0.420 | 0.300 | 0.415 | 0.579*** | 0.337 | 0.453* | 0.438* |  |  |  |  |
| Intent to use (L) | 0.425* | 0.149 | 0.323 | 0.277 | 0.365 | 0.481** | 0.334 | 0.236 | 0.429* | 0.522*** | 0.632*** |  |  |  |
| Replace human interaction (M) | 0.361 | -0.046 | 0.438* | 0.265 | 0.406 | 0.200 | 0.424* | 0.263 | 0.369 | 0.098 | 0.272 | 0.280 |  |  |
| Persuasive (N) | 0.393 | 0.104 | 0.541*** | 0.327 | 0.466* | 0.348 | 0.454* | 0.589*** | 0.249 | 0.461* | 0.353 | 0.323 | 0.336 |  |
| Education (O) | 0.030 | 0.044 | -0.174 | -0.044 | -0.251 | -0.210 | 0.043 | 0.087 | -0.175 | 0.113 | 0.232 | 0.173 | -0.180 | 0.086 |

\*Bayes Factor ( $BF_{10}$ ) > 10, \*\* $BF_{10}$ > 30, \*\*\* $BF_{10}$ > 100

### Quantitative findings using subset of the data who used ChatGPT for healthcare related queries

Correlation analyses

This sections reports finding based on 44 respondents who used ChatGPT for healthcare-related queries. Table 4 presents a nuanced landscape of significant correlations among various perceived attributes of ChatGPT as evaluated by users employing the AI for health-related inquiries. The significance of these correlations is underscored by BF_10_.

**Table 4.** Comparative Bayesian Analysis of Decision-Making Models (n=44).

| Models → Help make decision | $P(M)$ | $P(M\vert data)$ | $BF_M$ | $BF_{10}$ | $R^2$ |
| --- | --- | --- | --- | --- | --- |
| Trustworthy + Benefits outweigh risks + Intent to use + Education | $6.104 \times 10^{-5}$ | 0.006 | 106.540 | 1.000 | 0.617 |
| Trustworthy + ChatGPT for healthcare queries + Intent to use + Education | $6.104 \times 10^{-5}$ | 0.004 | 63.255 | 0.595 | 0.606 |
| Competent + Trustworthy + ChatGPT for healthcare queries | $6.104 \times 10^{-5}$ | 0.004 | 59.841 | 0.563 | 0.573 |
| Trustworthy + Benefits outweigh risks + Intent to use | $6.104 \times 10^{-5}$ | 0.004 | 59.065 | 0.556 | 0.572 |
| Competent + Trustworthy + ChatGPT for healthcare queries + Education | $6.104 \times 10^{-5}$ | 0.003 | 54.670 | 0.515 | 0.603 |
| Trustworthy + Benefits outweigh risks + Satisfaction + Intent to use + Education | $6.104 \times 10^{-5}$ | 0.003 | 51.690 | 0.478 | 0.631 |
| Competent + Trustworthy + ChatGPT for healthcare queries + Intent to use + Education | $6.104 \times 10^{-5}$ | 0.003 | 49.168 | 0.463 | 0.630 |
| Trustworthy + ChatGPT for healthcare queries + Benefits outweigh risks + Intent to use + Education | $6.104 \times 10^{-5}$ | 0.003 | 49.076 | 0.462 | 0.630 |
| Trustworthy + Benefits outweigh risks + Intent to use + Persuasive + Education | $6.104 \times 10^{-5}$ | 0.003 | 45.529 | 0.429 | 0.628 |
| Trustworthy + Intent to use + Education | $6.104 \times 10^{-5}$ | 0.003 | 43.929 | 0.414 | 0.565 |

$P(M)$ = prior probability of the model before observing the data (non-informative) $P(M|data)$ = posterior probability of the model given the observed data. $BF_M$ = Bayes Factor of the model relative to a baseline (null) model $F_{10}$ = Bayes Factor<sub>10</sub> refers to the comparison between the alternative hypothesis (H1) and the null hypothesis (H0)

A notable significant correlation (r=0.623***) exists between ‘Transparent’ and ‘Trustworthy’, suggesting a robust relationship where the clarity of ChatGPT’s processes and intentions is strongly associated with users’ trust. ‘Trustworthy’ also shares a substantial correlation (r=0.682***) with ‘Helps make decisions’, indicating that trust in ChatGPT is crucial for users considering its advice in making health-related decisions.

Additionally, ‘Secure’ exhibits very strong evidence of correlation (r=0.583***) with ‘ChatGPT for healthcare queries’, pointing to security as a pivotal factor for users when consulting ChatGPT for health concerns. The significant correlation between ‘Secure’ and ‘Not manipulate’ (r=0.583***) also highlights the interdependence of security and the non-manipulative nature of responses in fostering a safe environment for health-related interactions.

Furthermore, ‘Benefits outweigh risks’ has a very strong correlation (r=0.532***) with ‘ChatGPT for healthcare queries’, which could indicate that users who perceive higher benefits than risks are more likely to engage with ChatGPT for health-related purposes. ‘Intent to use’ shows a strong positive correlation (r=0.632***) with ‘Willing to take decisions’, suggesting that users who intend to use ChatGPT are also more likely to trust it for decision-making. The correlation of ‘Intent to use’ with ‘Satisfaction’ (r=0.522***) and ‘ChatGPT for healthcare queries’ (r=0.481**) emphasizes the connection between future use intentions, current satisfaction levels, and the perceived utility of ChatGPT in health-related matters. Lastly, ‘Persuasive’ (N) demonstrates very strong evidence of correlation (r=0.589***) with ‘ChatGPT for healthcare queries’, indicating that the ability of ChatGPT to influence user opinions or actions is significantly related to its use for health queries.

As illustrated in Figure 2, in the subsequent in-depth analysis of selected pairwise correlation, we investigated the relationship between ‘Help make decisions’ with ‘Competence,’ ‘Transparent,’ ‘Benefits outweigh risks,’ and ‘Persuasive’. We also explored pairwise correlation of ‘Trustworthy’ with ‘Transparent’ and ‘Persuasive’. Prior and posterior distributions for Pearson’s correlation coefficient were generated to elucidate the relationship between variables. A robustness check was conducted to ensure the stability of the Bayes Factor across varying priors. This analysis confirmed the strength of the evidence for the alternative hypothesis (H1) (see S1).

**Figure 2.** Bayesian Correlation Sequential Analyses of ChatGPT’s Attributes and Their Influence on Decision-Making Assistance. **A**: Correlation between perceived competence of ChatGPT and its assistance in decision-making, indicating very strong evidence for the positive association (BF_10_ = 86.0414); **B**: Association between perceived transparency of ChatGPT and its aid in decision-making, demonstrating strong evidence for the correlation (BF_10_ = 26.0618); **C**: Relationship between the perceived benefits outweighing risks when using ChatGPT for decision-making, showing strong evidence for the correlation (BF_10_ = 11.4741); **D**: Correlation between perceived persuasiveness of ChatGPT and its impact on decision-making, with anecdotal evidence for the association (BF_10_ = 0.677); **E**: Correlation between perceived trustworthiness and persuasiveness of ChatGPT, suggesting anecdotal evidence for their combined influence on decision-making assistance (BF_10_ = 0.544); **F**: Analysis of the relationship between transparency and trustworthiness in ChatGPT, with extreme evidence supporting a very strong correlation (BF_10_ = 3690).

The sequential analysis further reinforced our Table 4 findings. As additional data points were sequentially integrated into the analysis, the evidence for a positive correlation between the variable remained robustly within the threshold for extreme, very strong, and anecdotal evidence. This pattern persisted across the accumulation of data points, indicating that the observed correlation was not a consequence of sample size but a persistent trend within the data.

For the attribute of competence, the analysis indicated very strong evidence (BF_10_ = 86.0414) for the hypothesis that ChatGPT’s competence positively influences its ability to help users make decisions. This was further corroborated by the robustness checks, which consistently showed substantial support for this relationship across a spectrum of priors. The transparency of ChatGPT also appeared to be a significant factor, with a Bayes Factor suggesting strong evidence (BF_10_ = 26.0618) for its correlation with aiding decision-making. This aligns with the notion that transparency in the functioning of AI systems may bolster user trust and reliance on their decision-making capabilities.

In the case of the perceived benefits outweighing risks, there was strong evidence (BF_10_ = 11.4741) supporting the relationship with decision-making assistance. Users who felt that the advantages of using ChatGPT surpassed any potential risks were more likely to consider it helpful in making decisions. However, when examining the attribute of persuasiveness, the evidence was merely anecdotal (BF_10_ = 0.677), indicating a weak relationship with decision-making aid. This suggests that while ChatGPT’s ability to persuade may be noticed by users, it does not significantly influence their reliance on the system for decision-making support.

The trustworthiness attribute, when analyzed in conjunction with persuasiveness, showed a similarly modest level of evidence (BF_10_ = 0.544), again suggesting that these factors alone do not strongly predict ChatGPT’s perceived utility in decision-making. The most compelling result was observed in the correlation between transparency and trustworthiness, where an extreme Bayes Factor (BF_10_ = 3690) was noted, indicating an exceptionally strong relationship between these attributes. This underscores the integral role of transparent operations and trust in the perceived effectiveness of AI systems like ChatGPT in supporting user decisions. This conclusion aligns with the overarching narrative of the primary analysis and adds a layer of depth to the understanding of user perceptions of ChatGPT within the specific context of health-related inquiries.

#### Bayesian Linear Regression Analysis

Table 5 presents a Bayesian linear regression comparing various models to ascertain the factors influencing decision-making when ChatGPT is not used for healthcare-related queries. P(M) represents the prior probability of each model before data observation, assuming a non-informative or uniform prior. P(M|data) denotes the posterior probability, reflecting the model’s probability after considering the observed data. The Bayes Factor (BF_M_) and Bayes Factor_10_ (BF_10_) provide evidence strength for each model against a baseline model, with BF_M_ referencing the null model and BF_10_ comparing each alternative hypothesis to the null hypothesis. The coefficient of determination (R²) indicates the proportion of variance in decision-making that is predictable from the independent variables in each model. The models are ranked by the strength of evidence.

**Table 5.** Posterior Analysis of Predictors Influencing Decision-Making Using Bayesian Linear Regression (n=44).

| Coefficient | P(incl) | P(excl) | P<br>(incl\|data) | P<br>(excl\|data) | $BF_{inclusion}$ | Mean | Std.<br>Dev | 95% Credible<br>interval | |
| --- | --- | --- | --- | --- | --- | --- | --- | --- | --- |
|  |  |  |  |  |  |  |  | Lower | Upper |
| Intercept | 1.000 | 0.000 | 1.000 | 0.000 | 1.000 | 3.227 | 0.089 | 3.016 | 3.387 |
| Competent | 0.500 | 0.500 | 0.451 | 0.549 | 0.820 | 0.074 | 0.116 | -0.042 | 0.366 |
| Reliable | 0.500 | 0.500 | 0.270 | 0.730 | 0.371 | -0.018 | 0.079 | -0.247 | 0.121 |
| Transparent | 0.500 | 0.500 | 0.285 | 0.715 | 0.399 | -0.021 | 0.084 | -0.259 | 0.101 |
| Trustworthy | 0.500 | 0.500 | 0.988 | 0.012 | 82.394 | 0.454 | 0.137 | 0.156 | 0.684 |
| Not manipulate | 0.500 | 0.500 | 0.343 | 0.657 | 0.521 | 0.038 | 0.090 | -0.039 | 0.330 |
| Secure | 0.500 | 0.500 | 0.281 | 0.719 | 0.390 | 0.015 | 0.082 | -0.116 | 0.299 |
| ChatGPT for healthcare queries | 0.500 | 0.500 | 0.458 | 0.542 | 0.844 | 0.089 | 0.137 | -0.029 | 0.433 |
| Benefits outweigh risks | 0.500 | 0.500 | 0.516 | 0.484 | 1.065 | 0.097 | 0.129 | -0.048 | 0.381 |
| Satisfaction | 0.500 | 0.500 | 0.290 | 0.710 | 0.409 | -0.022 | 0.081 | -0.254 | 0.113 |
| Willing to take decisions | 0.500 | 0.500 | 0.297 | 0.703 | 0.423 | 0.019 | 0.080 | -0.080 | 0.247 |
| Intent to use | 0.500 | 0.500 | 0.554 | 0.446 | 1.245 | 0.119 | 0.144 | -0.020 | 0.453 |
| Replace human-to-human interaction | 0.500 | 0.500 | 0.295 | 0.705 | 0.419 | 0.024 | 0.079 | -0.094 | 0.261 |
| Persuasive | 0.500 | 0.500 | 0.306 | 0.694 | 0.440 | -0.031 | 0.099 | -0.343 | 0.088 |
| Education | 0.500 | 0.500 | 0.522 | 0.478 | 1.093 | -0.079 | 0.103 | -0.307 | 0.034 |

As shown in Table 5 the model comprising "Trustworthy + Benefits outweigh risks + Intent to use + Education" emerged as the most robust, reflected by the highest Bayes Factor in relation to the null model (BF_10_ = 106.540), suggesting substantial evidence in favor of this model compared to the alternative models considered. The coefficient of determination (R²) for this model was 0.617, indicating that approximately 61.7% of the variance in the use of ChatGPT for making decisions was accounted for by the predictors included in this model.

The posterior summaries (Table 6) of coefficients revealed that ‘Trustworthy’ had a posterior mean estimate of 0.454 with a 95% credible interval ranging from 0.156 to 0.684, indicating that as ChatGPT is perceived as more trustworthy, participants are more likely to use it for healthcare-related decision-making. P(incl) and P(excl) are the prior inclusion and exclusion probabilities, respectively, assuming an initial 50/50 split without evidence. P(incl|data) and P(excl|data) represent the posterior probabilities of inclusion and exclusion after data consideration. BF_inclusion_ stands for the Bayes Factor of inclusion, providing the evidence ratio for each predictor’s effect. The mean column represents the average posterior estimate of the effect size, while the standard deviation (Std. Dev) quantifies the uncertainty associated with the mean estimates. The 95% credible interval provides the range within which the true effect size is likely to fall with 95% certainty.

**Table 6.**
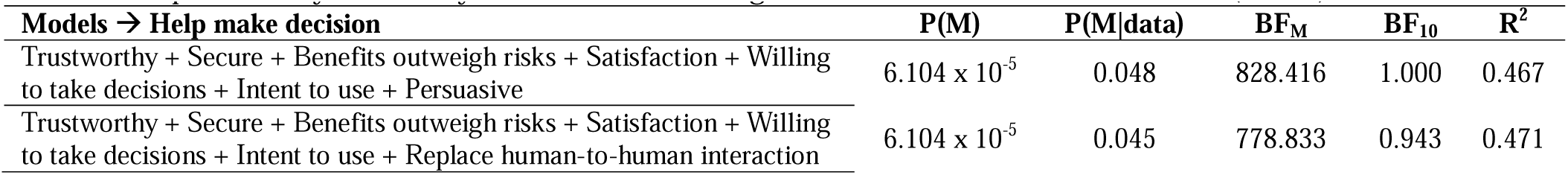
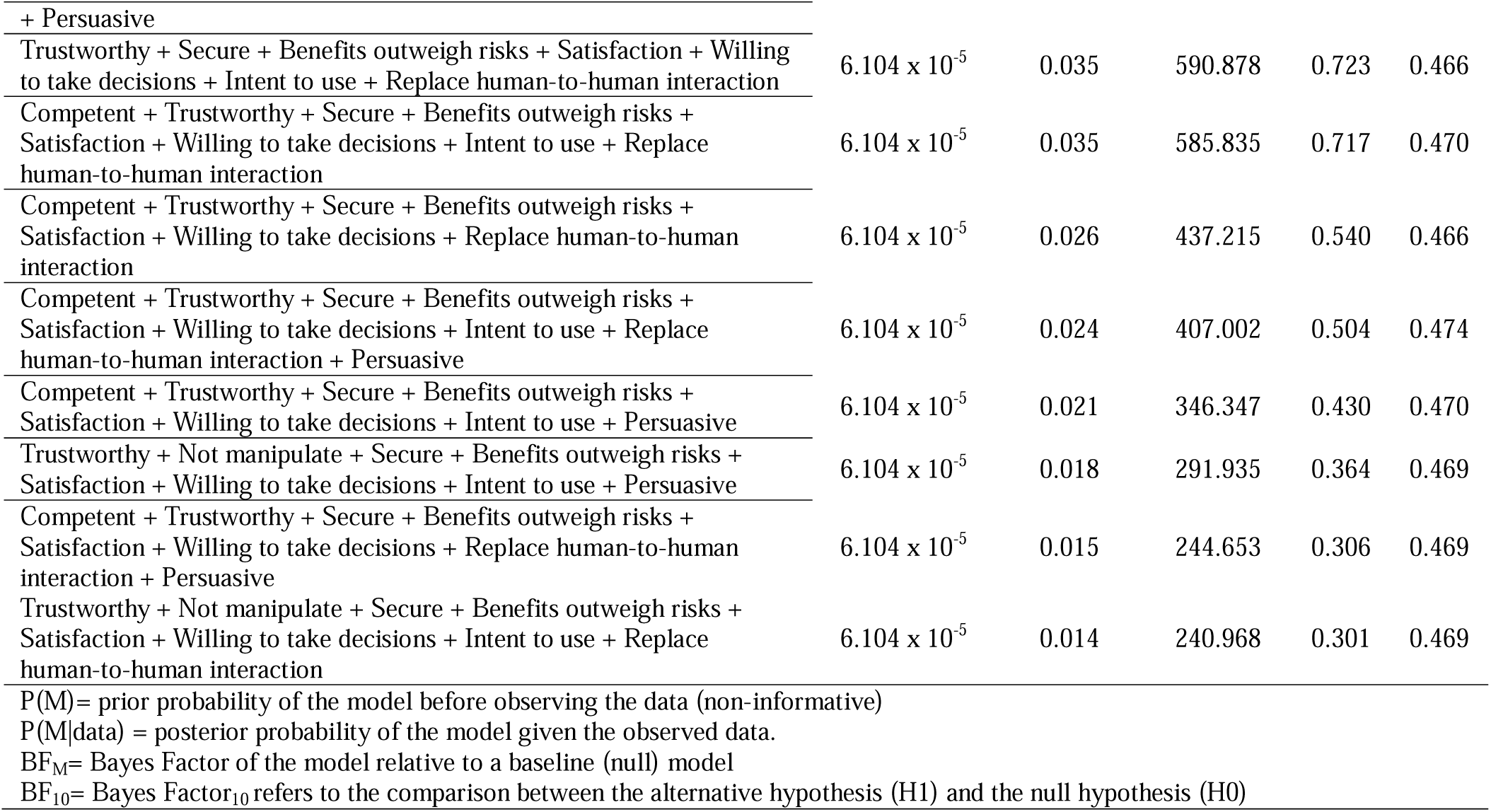
Comparative Bayesian Analysis of Decision-Making Models in Non-Healthcare Contexts (n=563).

### Quantitative findings using subset of the data who did not use ChatGPT for healthcare related queries

#### Bayesian correlation analysis

This sections reports finding based on 563 respondents who did not use ChatGPT for healthcare-related queries. The correlation analysis (see S2) demonstrated a complex interplay of attributes with certain pairs, such as ‘Competent’ and ‘Reliable’ or ‘Secure’ and ‘ChatGPT for healthcare queries’, showing very strong evidence of a positive relationship. These robust statistical associations imply that perceptions of competence, reliability, transparency, trustworthiness, and security are significantly interrelated with the use of ChatGPT for healthcare queries, the perceived benefits outweighing risks, and the ability of ChatGPT to help make decisions. Additionally, satisfaction with ChatGPT, willingness to take decisions based on ChatGPT’s advice, and intent to use ChatGPT in the future are also strongly interconnected, suggesting that positive user experience translates into continued use intentions.

#### Bayesian Linear Regression Analysis

Table 7 shows the model comparisons using BF_10_ identified the model with the highest evidence against the null model, consisting of the variables ‘Trustworthy’, ‘Secure’, ‘Benefits outweigh risks’, ‘Satisfaction’, ‘Willing to take decisions’, ‘Intent to use’, ‘Replace human-to-human interaction’, and ‘Persuasive’. This model achieved a BF_10_ of 828.416, suggesting very strong evidence in favor of this model over the null model, with an R² of 0.467, indicating it accounts for approximately 46.7% of the variance in the outcome.

**Table 7.** Posterior Analysis of Predictors Influencing Decision-Making Using Bayesian Linear Regression (n=563).

| Coefficient | P(incl) | P(excl) | P<br>(incl\|data) | P<br>(excl\|data) | BF<sub>inclusion</sub> | Mean | Std.<br>Dev | 95% Credible<br>interval |  |
| --- | --- | --- | --- | --- | --- | --- | --- | --- | --- |
|  |  |  |  |  |  |  |  | Lower | Upper |
| Intercept | 1.000 | 0.000 | 1.000 | 0.000 | 1.000 | 3.259 | 0.024 | 3.208 | 3.303 |
| Competent | 0.500 | 0.500 | 0.475 | 0.525 | 0.906 | 0.040 | 0.051 | 0.000 | 0.140 |
| Reliable | 0.500 | 0.500 | 0.124 | 0.876 | 0.142 | 0.002 | 0.016 | 0.000 | 0.059 |
| Transparent | 0.500 | 0.500 | 0.130 | 0.870 | 0.150 | 0.003 | 0.016 | -0.014 | 0.051 |
| Trustworthy | 0.500 | 0.500 | 0.783 | 0.217 | 3.606 | 0.077 | 0.052 | 0.000 | 0.156 |
| Not manipulate | 0.500 | 0.500 | 0.293 | 0.707 | 0.414 | 0.015 | 0.030 | 0.000 | 0.090 |
| Secure | 0.500 | 0.500 | 0.992 | 0.008 | 128.914 | 0.154 | 0.044 | 0.063 | 0.229 |
| ChatGPT for healthcare queries | 0.500 | 0.500 | 0.180 | 0.820 | 0.220 | 0.007 | 0.022 | -0.015 | 0.068 |
| Benefits outweigh risks | 0.500 | 0.500 | 0.995 | 0.005 | 209.320 | 0.163 | 0.044 | 0.080 | 0.247 |
| Satisfaction | 0.500 | 0.500 | 0.949 | 0.051 | 18.749 | 0.129 | 0.051 | 0.000 | 0.204 |
| Willing to take decisions | 0.500 | 0.500 | 0.986 | 0.014 | 69.047 | 0.141 | 0.043 | 0.062 | 0.233 |
| Intent to use | 0.500 | 0.500 | 0.739 | 0.261 | 2.838 | 0.076 | 0.057 | $-3.4 \times 10^{-4}$ | 0.165 |
| Replace human-to-human interaction | 0.500 | 0.500 | 0.666 | 0.334 | 1.995 | 0.050 | 0.044 | 0.000 | 0.122 |
| Persuasive | 0.500 | 0.500 | 0.598 | 0.402 | 1.487 | 0.055 | 0.055 | $-1.8 \times 10^{-4}$ | 0.154 |
| Education | 0.500 | 0.500 | 0.134 | 0.866 | 0.155 | 0.003 | 0.012 | -0.007 | 0.045 |

Table 8 summarizes the posterior distributions of regression coefficients for each variable contributing to decision-making efficacy. The posterior summaries of coefficients revealed that the credible intervals for ‘Secure,’ ‘Benefits outweigh risks,’ and ‘Willing to take decisions’ have a significant positive impact on the dependent variable (‘Help make decisions’). In other words, increases in these predictors are associated with an increase in the likelihood that ChatGPT helps users make informed and timely decisions.

#### Qualitative findings

Distinct patterns emerged when contrasting the concerns of users employing the ChatGPT for healthcare-related inquiries against those using it for other purposes. Both cohorts expressed concerns over privacy, a testament to the overarching necessity for robust data security protocols in AI interactions. However, the nuances of their feedback reveal divergent emphases reflective of the context of use.

Users engaging with ChatGPT for healthcare-related matters demonstrate a pronounced emphasis on safety and trust. This is indicative of the critical nature of healthcare information and the consequential outcomes dependent on its reliability. Concerns such as the AI becoming overly intelligent suggest apprehensions about the delegation of health-related decision-making to artificial entities, raising questions about the ethical bounds of AI in sensitive sectors. Moreover, while both sets of users highlight the importance of content quality, the healthcare user group uniquely underlines the dual dimensions of quantity and quality. This emphasis may underscore the necessity for comprehensive yet reliable health information that AI platforms like ChatGPT are expected to deliver. In contrast, the feedback from users not engaged in healthcare queries with ChatGPT tends to span a broader spectrum of technical and usability enhancements. Calls for a more user-friendly interface, and improved load times. Users also express the need for more human-like interactions, suggesting an aspiration for AI to bridge the gap between technological functionality and human relationality. Notably, users from the non-healthcare cohort voiced concerns about potential political or ideological biases in AI responses. This highlights a distinct concern for objectivity and neutrality, which, while important in all contexts, appears particularly salient for users seeking general information and assistance from ChatGPT.

The comparative analysis reveals that while the foundation of user trust in AI is built on privacy and accuracy, the specific context of use—healthcare versus general inquiries—exerts a significant influence on the nuances of user concerns. For healthcare-related AI applications, the implications are clear: there is a critical need for heightened measures of accuracy, security, and ethical considerations, aligning with the sensitive nature of healthcare information and decision-making processes.

## Discussion

In the realm of healthcare decision-making, the prominence of trustworthiness, as indicated by our study, is particularly instructive. Within our sample of 44 healthcare users of ChatGPT, trustworthiness emerged as a critical factor influencing their reliance on the platform for healthcare-related queries.

Firstly, the critical nature of healthcare decisions inherently demands a measured degree of trust. Decisions based on health-related information directly impact an individual’s physical well-being, making the accuracy and reliability of such information paramount. This is especially true in an era where digital health advice is becoming increasingly prevalent, and users often must navigate a plethora of information sources of varying credibility. In contrast, the non-healthcare user group, which is notably larger, might prioritize other factors like security, user experience, or the ability to supplement or replace human interaction. For these users, the stakes of the decision-making process, while still important, do not typically bear the direct and immediate implications for personal health and well-being. Consequently, their assessment of trustworthiness might be based more on factors like the platform’s ability to deliver satisfactory and efficient interactions.

Secondly, the complexity and validity of health information further elevates the need for trust. Healthcare information is often laden with medical jargon and intricate details about conditions and treatments, requiring users to place considerable trust in the source to ensure that the information is both understandable and accurate. Moreover, the personal and confidential nature of health information calls for a heightened level of trust in the platform’s ability to handle such data ethically and securely. That being said, it is often suggested to train GPT like ChatGPT on accurate, reliable, and perhaps patient health data making the AI suitable for personalized recommendations and conversations [24]. Although such discussions and recommendations are sound, they hold a critical latent risk. As recently brought to notice, large language and GPT models tend to memorize training data, which can lead to privacy concerns [14]. If ChatGPT or similar AI models are trained on sensitive healthcare data in the future to provide personalized medical information, (with the hope of improving user experience and trust) this could introduce a significant risk of large-scale data breaches [25, 26]. Such a situation could have far-reaching consequences, as the breach of healthcare data not only compromises individual privacy but also violates legal and ethical standards in medical confidentiality. It could lead to misuse of personal health information, identity theft, and a loss of public trust in healthcare technology. Therefore, while the advancement of AI in healthcare promises significant benefits, including personalized and efficient patient care, it also necessitates robust security measures and ethical considerations to protect sensitive patient data against potential breaches.

The recent discovery of ChatGPT’s limitation [14] calls for urgent discussion about Technology Readiness Levels (TRLs) [27]. TRLs are a systematic metric used to assess the development stage of a technology, ranging from the conceptual stage (TRL 1) to full-scale deployment (TRL 9) [28]. This metric is pivotal in determining whether a technology is sufficiently mature for public use, particularly when it involves sensitive data. ChatGPT demonstrates immense potential in transforming healthcare through personalized medicine and efficient data processing. However, the integration of AI in healthcare necessitates a cautious approach, underscored by the rigorous assessment of TRLs. AI systems dealing with healthcare data must not only demonstrate advanced functionality but also robust security protocols to protect sensitive patient information. Assessing AI technologies against high TRL standards ensures that they have undergone extensive testing and validation in real-world scenarios, which includes evaluating their resilience against data breaches and extraction attacks.

Moreover, the importance of reaching higher TRLs before public deployment cannot be overstated in the context of public trust and safety. Prematurely launched AI technologies that have not been thoroughly vetted for security risks can lead to significant breaches of patient privacy. Such incidents not only erode public confidence in AI healthcare applications but also pose severe ethical and legal challenges. The healthcare sector, bound by stringent regulations and ethical standards, requires that any deployed technology adheres to the highest levels of confidentiality and data protection.

In enhancing and extending the discussion on the limitations of our study, it’s crucial to address both the methodological constraints and the statistical considerations of our approach. The discrepancy in sample sizes between the healthcare user group (n=44) and the non-healthcare user group (n=563) is a limitation. We advocate for a cautious interpretation of the results, particularly in applying our findings to a broader population of healthcare information seekers. Moreover, our sampling methodology, primarily through social media channels, might introduce a selection bias. The demographic that engages with healthcare-related AI through social media platforms may not accurately represent the broader population seeking healthcare information. Reliance on self-reported data is another methodological constraint. While self-reporting is a common and valuable tool in research, it inherently carries the risk of inaccuracies. Participants’ responses could be influenced by their perceptions, experiences, or even social desirability bias, which might affect the authenticity of the data. To mitigate some of these limitations, our study employed a Bayesian analytical approach. The Bayesian method is advantageous in dealing with smaller sample sizes [29]. Despite these methodological efforts, further research with a more diverse and larger sample, perhaps supplemented with objective usage metrics, would be beneficial to validate and expand our findings and to develop a more comprehensive understanding of how different user groups perceive and use AI in healthcare contexts.

In conclusion our study findings suggest a clear demarcation in user expectations and requirements from AI systems based on the context of their use. In healthcare, where decisions have direct health implications, trustworthiness emerges as a paramount concern, emphasizing the need for AI systems that prioritize accuracy, clarity, and security. For non-healthcare contexts, while trustworthiness remains important, it encompasses a broader range of factors including efficiency, user experience, and novelty. This dichotomy provides valuable insights for AI developers and healthcare professionals. It suggests a tailored approach to AI system design and communication, one that is acutely sensitive to the context of use. Future research might focus on exploring these context-specific preferences in greater detail, potentially leading to more personalized and effective AI applications across various domains.

## Data Availability

Anonymized data are available on personal contact with the corresponding author. Due to privacy concerns the raw data not available on any public repository.

## Supporting information

**S1. Supplementary file.** Extra analysis and results

**S2. Supplementary file.** Correlation analysis

